# Place-based disorder, social cohesion, and physical and mental health outcomes of LGBTQIA+ adults in the US

**DOI:** 10.1101/2025.09.04.25334946

**Authors:** Nguyen K. Tran, David H. Rehkopf, Charleigh Flohr, Juno Obedin-Maliver, Annesa Flentje, Mitchell R. Lunn

## Abstract

Place-based disorder and social cohesion may influence LGBTQIA+ adults’ physical and mental health in meaningful ways given their heightened exposure to minority stress and discrimination. However, few studies have examined these associations. Using a sample of 3790 LGBTQIA+ adults in 786 counties from The Population Research in Identity and Disparities for Equality Study, we assessed associations of place-based disorder and social cohesion with depressive symptoms, perceived stress, and physical health using linear mixed models. We tested effect modification by gender modality and ethnoracial group. Residing in counties with higher social cohesion by 1-unit was associated with 1.06 lower points (95% CI -1.56, -0.56) for depressive symptoms, 1.60 lower points (95% CI -2.26, -0.94) for perceived stress, and 1.17 higher points (95% CI 0.43, 1.90) for physical health. Residence in counties with higher place-based disorder by 1-unit was only associated with 1.17 higher points (95% CI 0.32, 2.01) for perceived stress; no association was observed for depressive symptoms or physical health. Findings indicate that physical and social environments are important to the health of LGBTQIA+ individuals.

## INTRODUCTION

Lesbian, gay, bisexual, transgender, queer, intersex, asexual, and other sexual and gender minority (LGBTQIA+) adults experience notable inequities in physical and mental health.^1–9^ Compared to their non-LGBTQIA+ peers, LGBTQIA+ adults report higher levels of anxiety, depression, substance use, and suicide attempts.^2–4,7,8^ Physical health disparities have also been documented, including elevated prevalences of asthma, cancer, cardiovascular disease, dementia, obesity, and stroke.^1,2,5–7^ These inequities are often explained using the minority stress framework, which attributes poor health outcomes to the additional stress stemming from discrimination, stigma, and social exclusion specific to one’s marginalized identity.^10–12^ Prior studies indicate that factors such as anti-transgender policies, conversion practices (*i*.*e*., organized attempts that seek to suppressed noncisgender gender identity and nonheterosexual sexual attraction), LGBTQIA+-targeted violence, unsafe environments, discrimination experiences, and internalized stigma were associated with poorer mental and physical health outcomes among LGBTQIA+ individuals.^13–15^

*Healthy People 2030* represents the latest national effort to promote health and prevent disease in the US by targeting key social determinants of health.^16^ Among its central priorities are improving the physical and social environments, which shape a wide range of health outcomes. In alignment with these goals, a growing body of research has examined the relationship between area characteristics (*i*.*e*., the physical, social, and economic features of the places where people live) and health outcomes among LGBTQIA+ populations.^17–22^ Two important features of the physical and social environment shown to be associated with health outcomes among LGBTQIA+ adults are place-based disorder and social cohesion. Place-based disorder, which includes indicators such as physical neglect, violence, or instability,^23^ has been associated with elevated levels psychological distress, substance use, and inflammatory biomarkers.^18,19,22^ Conversely, social cohesion reflects mutual trust, shared norms, and community connectedness,^24,25^ and has been associated with a lower prevalence of severe psychological distress and HIV-related sexual behaviors.^17,21^ These area characteristics may operate through multiple pathways, including exposure to stress-related processes, changes in health behaviors, access to resources, and perceptions of safety or belonging that ultimately contribute to health and health inequities.^26^ Importantly, the health-related impact of area-level conditions is unevenly distributed across populations with Black residents disproportionately affected by area-level disadvantage due to residential segregation.^27–29^

A 2022 review indicated that empirical evidence on place-based disorder, and residential environments more broadly, among LGBTQIA+ individuals remains limited,^30^ with current studies constrained by various limitations. First, the extant research has predominately sampled participants from urban settings^17–19,21,22^ that may not reflect the experiences of LGBTQIA+ individuals more broadly, as features of the physical and social environment may differ substantially across geographic areas. Second, most studies have been limited in sample size,^17–22^ which increases uncertainty of the estimated association due to sampling error and makes it challenging to identify LGBTQIA+ subgroup differences. Third, prior studies are often situated within the context of HIV prevention.^17–19,22^ This focus overlooks the broader range of physical and mental health outcomes that have been documented to disproportionately affect LGBTQIA+ individuals. Thus, studies with larger samples across diverse geographic settings and research context is needed to unsderstand how place-based disorder and social cohesion contribute to health inequities among LGBTQIA+ individuals for planning place-based interventions that can address these upstream, contextual determinants.

Our study addresses these gaps by examining associations between place-based disorder and social cohesion with physical and mental health outcomes in a sample of LGBTQIA+ adults. We further test whether these associations differ by ethnoracial group and gender modality to better understand how racial discrimination and cissexism may shape the ways participants access and afford to live in certain areas, thus modifying the relationship between area-level characteristics and individual health.

## METHODS

### Study design and participants

We conducted a cross-sectional study using data from The Population Research in Identity and Disparities for Equality (PRIDE) Study — a national, online, longitudinal cohort of LGBTQIA+ adults in the United States. Details of the study design and community engagement efforts have been described previously.^31,32^ Briefly, the cohort includes self-identified LGBTQIA+ adults aged 18 years or older who reside in the United States or its territories and are able to read and understand English. Recruitment began in May 2017 through social media, digital advertisements, and outreach at LGBTQIA+ community events and organizations. Upon enrollment, participants were invited to complete annual health questionnaires generally administered from June/July to May/June of the following year.

The current analysis used data from a subset of participants who completed a one-time “Social Determinants of Health” ancillary questionnaire administered from March to June 2023. The questionnaire, which as developed based on the *All of Us* Research Program’s survey instrument,^33^ assessed area characteristics, social relationships, stress, and perceptions of daily life. We linked and harmonized individual-level data from this questionnaire to participants’ (i) most recently completed annual health questionnaire from 2021 to 2023, (ii) home address reported through The PRIDE Study’s participant portal, and (iii) county-level socioeconomic conditions from the American Community Survey. We used counties as the administrative unit of analysis, as they may be relevant for understanding features of the physical and social environment while ensuring that a sufficient number of counties had multiple residents.^34^ Participants were included in the analytic sample if they completed the “Social Determinants of Health” questionnaire, at least one annual health questionnaire between 2021 and 2023, and provided a valid home address (**Figure S1**). Those with missing data on area characteristics, health outcomes, gender identity, and ethnoracial identity were excluded from the sample. Characteristics of included and excluded participants with linked annual questionnaire data are presented in **Table S1**.

The PRIDE Study is approved by the WIRB-Copernicus Group and Stanford University institutional review boards. All participants provided electronic informed consent.

### Measures

The primary outcomes were mean levels of depressive symptoms, perceived stress, and general physical health. Depressive symptoms in the past 2 weeks were measured in the annual health questionnaire using the 9-item Patient Health Questionnaire (PHQ-9) scale.^35^ Responses were rated on a 4-point scale (0=not at all, to 3=nearly every day), and total scores ranged from 0 to 27 with higher scores indicating more symptoms (Cronbach’s α=0.89). Perceived stress was measured in the Social Determinants of Health questionnaire using the 10-item Cohen’s Perceived Stress Scale (PSS-10), an instrument designed to assess the degree to which situations in one’s life are appraised as stressful.^36^ Items assess how often respondents experienced stress-related feelings over the past month (*e*.*g*., feeling nervous or unable to control important things) using a 5-point scale (0=never to 4=very often). Scores were calculated by reverse-scoring positively worded items and summing all items, with higher scores indicating greater perceived stress (Cronbach’s α=0.93). General physical health was assessed in the annual health questionnaire using the Global Physical Health (GPH) subscale of the Patient-Reported Outcomes Measurement Information System (PROMIS) Global Health measure.^37^ The GPH score was derived from 4 items assessing general physical health, physical functioning, pain, and fatigue. Each item, aside from pain, was rated on a 5-point Likert scale, with higher values representing better health. Pain was rescored to a 5-point scale as recommend (0 = 5; 1-3 = 4; 4-6 = 3; 7-9 = 2; 10 = 1) with higher values representing less pain. Raw scores were calculated (range=4-20) and standardized to a T-score based on the US general population (mean=50, SD=10).

Two area characteristics from the Social Determinants of Health questionnaire were examined: (i) place-based disorder and (ii) social cohesion (**Table S2**). These constructs reflect related but distinct social processes that are shaped by area-based socioeconomic conditions and have important implications for health outcomes.^38^ Place-based disorder was assessed using the 13-item Ross and Mirowsky Neighborhood Physical and Social Disorder Scale.^23^ Items were rated on a 4-point Likert scale and measured negative physical conditions (*e*.*g*., graffiti, excessive noise, vandalism, abandoned buildings) and psychosocial concerns (*e*.*g*., perceived safety, conflicts with neighbors). Positively worded items were reverse coded, and mean scores were calculated with higher scores indicating greater perceived disorder. Social cohesion was measured using 4 items from a validated scale assessing agreement on whether neighbors willingly help each other, get along, can be trusted, and share the same values, each rated on a 5-point Likert scale.^25^ Mean scores were computed with higher values indicating greater social cohesion. Both scales demonstrated good internal consistency (place-based disorder Cronbach’s α=0.91; social cohesion Cronbach’s α=0.81).

To obtain area-level measures of disorder and social cohesion for each county, we used a similar approach outlined in prior works^38^ to aggregate each participant’s disorder and social cohesion scores by calculating the mean of other participants’ responses residing in the same county. For counties with only one resident, the original score was retained. By aggregating responses from multiple participants in a given geographic area, this approach reduces measurement error from individual subjectivities and the potential for same source bias.^25^

Individual-level socio-demographics included age, gender identity, sexual orientation, ethnoracial identity, education level, and household income. Participants could select multiple ethnoracial identities. Based on these responses, individuals were classified as either belonging to at least 1 ethnoracial minority group or as white only.^39^ Gender modality was determined by cross-tabulating gender identity and sex assigned at birth to identify cisgender and transgender or gender diverse participants.^40,41^ County-level variables, including the proportion of adults aged 25 and older with at least a high school diploma and the proportion living below the federal poverty level, were derived from the 2022 American Community Survey 5-year estimates and linked to participants’ county of residence using geocoded home addresses.

### Statistical analysis

We conducted our analysis in four steps. First, we described the individual-level sociodemographic characteristics and county-level socioeconomic conditions for the overall sample and by place-based disorder and social cohesion. For this descriptive analysis, we dichotomized place-based disorder and social cohesion scores to be above and below the median. Differences were assessed using t-test or Mann-Whitney U-test for continuous variables and chi-square or Fisher’s Exact test for categorical variables. We calculated Pearson correlation coefficients between place-based disorder, social cohesion, and couny-level socioeconomic conditions to evaluate whether these area characteristics captured some components of the objective realities of participants’ county of residence. Second, we estimated the between-county variability in the outcomes by fitting a two-level null linear mixed model with individuals nested within counties and calculated the intraclass correlation coefficient to determine the degree of clustering for each outcome. Third, to assess the associations between place-based disorder and social cohesion with individual health outcomes, we fitted a series of two-level random-intercept linear mixed models. Due to the small amount of missing data (1% of participants), we pursued complete case analysis. Model 1 included place-based disorder and social cohesion separately as continuous variables to examine the unadjusted associations. Models 2-4 introduced potential confounders informed by a directed acyclic diagram (**Figure S2**) to account for variables identified from published literature that affected the distributions of place-based disorder and social cohesion. Model 2 added age (years) and its quadratic term, and Model 3 added ethnoracial group, education level, gender modality, and household income to adjust for demographic characteristics and adulthood socioeconomic position that may influence differential selection of LGBTIQA+ participants into counties with different levels of place-based disorder and social cohesion. To adjust for county-level socioeconomic conditions related to economic distress within counties, Model 4 added the county-level proportion of adults aged 25 and older with a high school diploma and the proportion of those living below the poverty level. Fourth, we explored effect modification by testing interaction terms between each area-level characteristic by ethnoracial group and by gender modality. All analyses were conducted in R version 4.5.1 with mixed models fitted using *lme4*.^42^

In sensitivity analysis, we used unconditional empirical Bayes estimates of area-level measures for each county to account for small within-county sample sizes that may contribute to measurement error.^43^ Empirical Bayes estimates, which is a weighted average of the mean for each county and the mean across all counties, were derived using a three-level linear mixed model that accounted for scale items nested in participants who are nested in counties. The weights are prorportional to the reliability of the area-level measure and result in greater shrinkage of counties with poorer reliability towards the overall mean, thus improving estimation for counties with small sample sizes.

## RESULTS

The analytic sample included 3790 participants from 786 counties. The number of participants in each county ranged from one to 114, and 418 counties had two or more residents (**Table S3**). The median age was 37.2 years (interquartile range=29.4-54.0; **Table 1**). Overall, 1741 (45.9%) were transgender or gender diverse, 695 (18.3%) reported at least 1 minoritized ethnoracial identity, 1593 (42.0%) identified as gay or lesbian, 2907 (76.7%) completed at least a 4-year degree, and 2409 (63.6%) had a household income of more than $50,000.

**Table 1.**
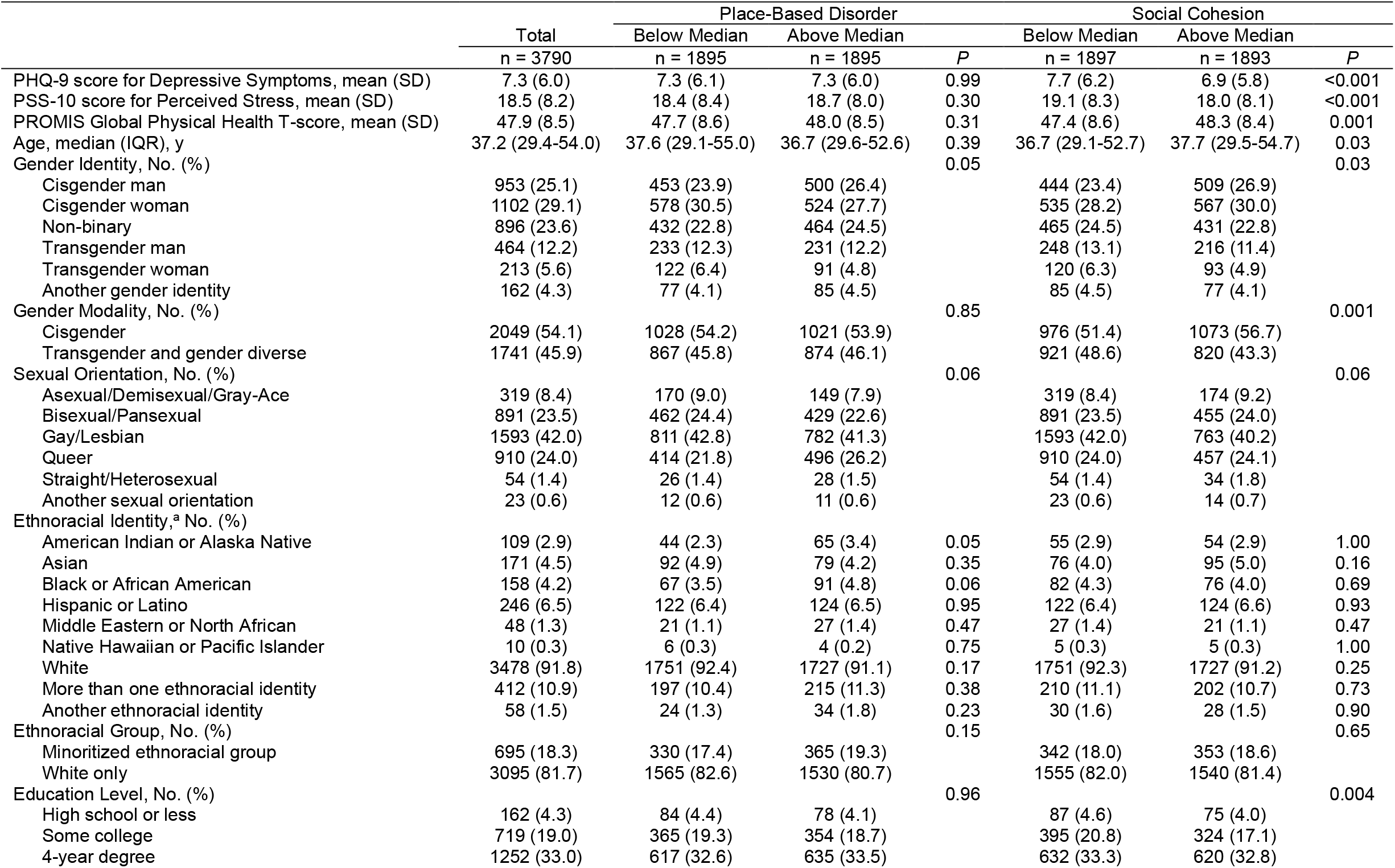
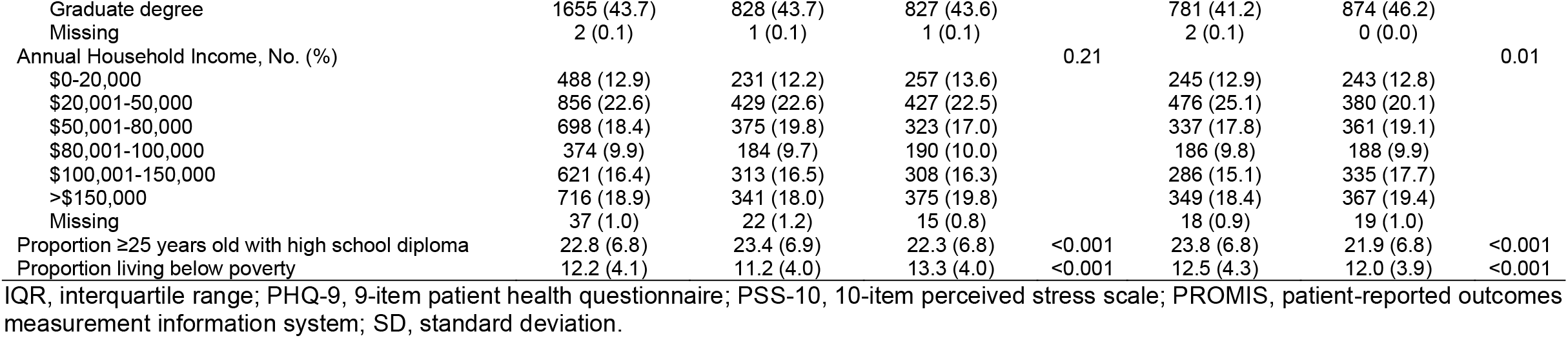
Individual health outcomes, sociodemographic factors, county-level socioeconomic conditions by area-level characteristics among LGBTQIA+ adults in The PRIDE Study.

|  | Total | Place-Based Disorder |  | P | Social Cohesion |  | P |
| --- | --- | --- | --- | --- | --- | --- | --- |
|  |  | Below Median | Above Median |  | Below Median | Above Median |  |
|  | n = 3790 | n = 1895 | n = 1895 |  | n = 1897 | n = 1893 |  |
| PHQ-9 score for Depressive Symptoms, mean (SD) | 7.3 (6.0) | 7.3 (6.1) | 7.3 (6.0) | 0.99 | 7.7 (6.2) | 6.9 (5.8) | <0.001 |
| PSS-10 score for Perceived Stress, mean (SD) | 18.5 (8.2) | 18.4 (8.4) | 18.7 (8.0) | 0.30 | 19.1 (8.3) | 18.0 (8.1) | <0.001 |
| PROMIS Global Physical Health T-score, mean (SD) | 47.9 (8.5) | 47.7 (8.6) | 48.0 (8.5) | 0.31 | 47.4 (8.6) | 48.3 (8.4) | 0.001 |
| Age, median (IQR), y | 37.2 (29.4-54.0) | 37.6 (29.1-55.0) | 36.7 (29.6-52.6) | 0.39 | 36.7 (29.1-52.7) | 37.7 (29.5-54.7) | 0.03 |
| Gender Identity, No. (%) |  |  |  | 0.05 |  |  | 0.03 |
| Cisgender man | 953 (25.1) | 453 (23.9) | 500 (26.4) |  | 444 (23.4) | 509 (26.9) |  |
| Cisgender woman | 1102 (29.1) | 578 (30.5) | 524 (27.7) |  | 535 (28.2) | 567 (30.0) |  |
| Non-binary | 896 (23.6) | 432 (22.8) | 464 (24.5) |  | 465 (24.5) | 431 (22.8) |  |
| Transgender man | 464 (12.2) | 233 (12.3) | 231 (12.2) |  | 248 (13.1) | 216 (11.4) |  |
| Transgender woman | 213 (5.6) | 122 (6.4) | 91 (4.8) |  | 120 (6.3) | 93 (4.9) |  |
| Another gender identity | 162 (4.3) | 77 (4.1) | 85 (4.5) |  | 85 (4.5) | 77 (4.1) |  |
| Gender Modality, No. (%) |  |  |  | 0.85 |  |  | 0.001 |
| Cisgender | 2049 (54.1) | 1028 (54.2) | 1021 (53.9) |  | 976 (51.4) | 1073 (56.7) |  |
| Transgender and gender diverse | 1741 (45.9) | 867 (45.8) | 874 (46.1) |  | 921 (48.6) | 820 (43.3) |  |
| Sexual Orientation, No. (%) |  |  |  | 0.06 |  |  | 0.06 |
| Asexual/Demisexual/Gray-Ace | 319 (8.4) | 170 (9.0) | 149 (7.9) |  | 319 (8.4) | 174 (9.2) |  |
| Bisexual/Pansexual | 891 (23.5) | 462 (24.4) | 429 (22.6) |  | 891 (23.5) | 455 (24.0) |  |
| Gay/Lesbian | 1593 (42.0) | 811 (42.8) | 782 (41.3) |  | 1593 (42.0) | 763 (40.2) |  |
| Queer | 910 (24.0) | 414 (21.8) | 496 (26.2) |  | 910 (24.0) | 457 (24.1) |  |
| Straight/Heterosexual | 54 (1.4) | 26 (1.4) | 28 (1.5) |  | 54 (1.4) | 34 (1.8) |  |
| Another sexual orientation | 23 (0.6) | 12 (0.6) | 11 (0.6) |  | 23 (0.6) | 14 (0.7) |  |
| Ethnoracial Identity,<sup>a</sup> No. (%) |  |  |  |  |  |  |  |
| American Indian or Alaska Native | 109 (2.9) | 44 (2.3) | 65 (3.4) | 0.05 | 55 (2.9) | 54 (2.9) | 1.00 |
| Asian | 171 (4.5) | 92 (4.9) | 79 (4.2) | 0.35 | 76 (4.0) | 95 (5.0) | 0.16 |
| Black or African American | 158 (4.2) | 67 (3.5) | 91 (4.8) | 0.06 | 82 (4.3) | 76 (4.0) | 0.69 |
| Hispanic or Latino | 246 (6.5) | 122 (6.4) | 124 (6.5) | 0.95 | 122 (6.4) | 124 (6.6) | 0.93 |
| Middle Eastern or North African | 48 (1.3) | 21 (1.1) | 27 (1.4) | 0.47 | 27 (1.4) | 21 (1.1) | 0.47 |
| Native Hawaiian or Pacific Islander | 10 (0.3) | 6 (0.3) | 4 (0.2) | 0.75 | 5 (0.3) | 5 (0.3) | 1.00 |
| White | 3478 (91.8) | 1751 (92.4) | 1727 (91.1) | 0.17 | 1751 (92.3) | 1727 (91.2) | 0.25 |
| More than one ethnoracial identity | 412 (10.9) | 197 (10.4) | 215 (11.3) | 0.38 | 210 (11.1) | 202 (10.7) | 0.73 |
| Another ethnoracial identity | 58 (1.5) | 24 (1.3) | 34 (1.8) | 0.23 | 30 (1.6) | 28 (1.5) | 0.90 |
| Ethnoracial Group, No. (%) |  |  |  | 0.15 |  |  | 0.65 |
| Minoritized ethnoracial group | 695 (18.3) | 330 (17.4) | 365 (19.3) |  | 342 (18.0) | 353 (18.6) |  |
| White only | 3095 (81.7) | 1565 (82.6) | 1530 (80.7) |  | 1555 (82.0) | 1540 (81.4) |  |
| Education Level, No. (%) |  |  |  | 0.96 |  |  | 0.004 |
| High school or less | 162 (4.3) | 84 (4.4) | 78 (4.1) |  | 87 (4.6) | 75 (4.0) |  |
| Some college | 719 (19.0) | 365 (19.3) | 354 (18.7) |  | 395 (20.8) | 324 (17.1) |  |
| 4-year degree | 1252 (33.0) | 617 (32.6) | 635 (33.5) |  | 632 (33.3) | 620 (32.8) |  |
| Graduate degree | 1655 (43.7) | 828 (43.7) | 827 (43.6) |  | 781 (41.2) | 874 (46.2) |  |
| Missing | 2 (0.1) | 1 (0.1) | 1 (0.1) |  | 2 (0.1) | 0 (0.0) |  |
| Annual Household Income, No. (%) |  |  |  | 0.21 |  |  | 0.01 |
| \$0-20,000 | 488 (12.9) | 231 (12.2) | 257 (13.6) | | 245 (12.9) | 243 (12.8) | |
| \$20,001-50,000 | 856 (22.6) | 429 (22.6) | 427 (22.5) | | 476 (25.1) | 380 (20.1) | |
| \$50,001-80,000 | 698 (18.4) | 375 (19.8) | 323 (17.0) | | 337 (17.8) | 361 (19.1) | |
| \$80,001-100,000 | 374 (9.9) | 184 (9.7) | 190 (10.0) | | 186 (9.8) | 188 (9.9) | |
| \$100,001-150,000 | 621 (16.4) | 313 (16.5) | 308 (16.3) | | 286 (15.1) | 335 (17.7) | |
| >\$150,000 | 716 (18.9) | 341 (18.0) | 375 (19.8) | | 349 (18.4) | 367 (19.4) | |
| Missing | 37 (1.0) | 22 (1.2) | 15 (0.8) |  | 18 (0.9) | 19 (1.0) |  |
| Proportion ≥25 years old with high school diploma | 22.8 (6.8) | 23.4 (6.9) | 22.3 (6.8) | <0.001 | 23.8 (6.8) | 21.9 (6.8) | <0.001 |
| Proportion living below poverty | 12.2 (4.1) | 11.2 (4.0) | 13.3 (4.0) | <0.001 | 12.5 (4.3) | 12.0 (3.9) | <0.001 |

IQR, interquartile range; PHQ-9, 9-item patient health questionnaire; PSS-10, 10-item perceived stress scale; PROMIS, patient-reported outcomes measurement information system; SD, standard deviation.
<sup>a</sup> Participants were able to select multiple options, thus, percentages do not equal to 100%.

Mean scores were 1.81 (median=1.84, standard deviation [SD]=0.29) for place-based disorder and 3.45 (median=3.49, SD=0.36) for social cohesion. A higher proportion of cisgender men, non-binary participants, American Indian or Alaska Natives, and Black or African Americans resided in counties with place-based disorder scores above the median (**Table 1**). Those who were younger, identified as transgender and gender diverse, without a 4-degree, and had a household income ≤$50,000 were more likely to resided in counties with social cohesion scores below the median. Place-based disorder and social cohesion appeared to have captured different components of county-level socioeconomic conditions (**Figure S3**). Place-based disorder was weakly correlated with the proportion living below poverty (*r*=0.24), but not with the proportion ≥25 years old with at least a high school diploma (*r*= -0.03). Social cohesion was weakly correlated with the proportion ≥25 years old with at least a high school diploma (*r*= - 0.19) but not with proportion living below poverty (*r*= -0.05).

Mean scores were 7.3 (SD=6.0) for PHQ-9, 18.5 (SD=8.2) for PSS-10, and 47.9 (SD=8.5) for GPH T-score (**Table 1**). Participants residing in counties with social cohesion scores above the median had lower individual scores for depressive symptoms (*P*<0.001), perceived stress (*P*<0.001), and higher T-scores for physical health (*P*=0.001). No differences were observed for place-based disorder. In the null mixed effects model, although most of the variability in each outcome occurred at the individual level, a modest proportion was accounted for at the county level for depressive symptoms (4.0%), perceived stress (2.7%), and physical health (4.0%; **Table S4**).

In unadjusted linear mixed models, participants who resided in counties with higher levels of social cohesion had fewer depressive symptoms, lower perceived stress, and better physical health (**Table 2, Figure 1**). These associations persisted after adjusting for individual and county-level confounders, with a 1-unit higher score in social cohesion associated with a PHQ-9 score that is 1.06 points lower (95% confidence interval [CI] -1.56, -0.56), a PSS-10 score that is 1.60 points lower (95% CI -2.26, -0.94), and a GPH T-socre that is 1.17 points higher (95% CI 0.42, 1.90). For place-based disorder, higher scores were associated with poorer outcomes across all three measures in unadjusted models (**Table 2, Figure 2**). After adjusting for individual- and county-level confounders, associations persisted only for perceived stress, with a 1-unit higher score in place-based disorder associated with a PSS-10 score that is 1.17 points higher (95% CI 0.32, 2.01).

**Table 2.** Associations of area-level characteristics and individual-level depressive symptoms, perceived stress, and general physical health among LGBTQIA+ adults, The PRIDE Study.

|  | No.<br>Individuals | No.<br>Counties | Place-Based Disorder |  | Social Cohesion |  |
| --- | --- | --- | --- | --- | --- | --- |
|  |  |  | *B* (95% CI) | *P* | *B* (95% CI) | *P* |
| ***PHQ-9 scores for Depressive Symptoms*** |  |  |  |  |  |  |
| Model 1 | 3752 | 778 | 1.12 (0.38, 1.86) | 0.003 | -1.80 (-2.35, -1.25) | <0.001 |
| Model 2 | 3752 | 778 | 0.72 (0.01, 1.42) | 0.05 | -1.50 (-2.03, -0.98) | <0.001 |
| Model 3 | 3752 | 778 | 0.36 (-0.27, 0.99) | 0.27 | -1.19 (-1.68, -0.70) | <0.001 |
| Model 4 | 3752 | 778 | 0.52 (-0.13, 1.17) | 0.11 | -1.06 (-1.56, -0.56) | <0.001 |
| ***PSS-10 scores for Perceived Stress*** |  |  |  |  |  |  |
| Model 1 | 3752 | 778 | 2.29 (1.30, 3.28) | <0.001 | -2.65 (-3.39, -1.90) | <0.001 |
| Model 2 | 3752 | 778 | 1.51 (0.61, 2.41) | 0.001 | -2.16 (-2.84, -1.48) | <0.001 |
| Model 3 | 3752 | 778 | 1.02 (0.21, 1.84) | 0.02 | -1.68 (-2.33, -1.03) | <0.001 |
| Model 4 | 3752 | 778 | 1.17 (0.32, 2.01) | 0.007 | -1.60 (-2.26, -0.94) | <0.001 |
| ***PROMIS Global Physical Health T-score*** |  |  |  |  |  |  |
| Model 1 | 3752 | 778 | -1.10 (-2.16, -0.05) | 0.04 | 1.93 (1.14, 2.72) | <0.001 |
| Model 2 | 3752 | 778 | -0.90 (-1.96, 0.15) | 0.09 | 1.77 (0.99, 2.56) | <0.001 |
| Model 3 | 3752 | 778 | -0.35 (-1.30, 0.60) | 0.47 | 1.27 (0.54, 2.00) | 0.001 |
| Model 4 | 3752 | 778 | -0.56 (-1.53, 0.40) | 0.25 | 1.17 (0.43, 1.90) | 0.002 |

Models 1 included each area characteristic in a separate model. Model 2 added mean centered age and its quadratic term. Model 3 added ethnoracial group, education level, gender modality, and household income. Model 4 added proportion $\geq 25$ years old with high school diploma (or equivalency) and proportion living below federal poverty level. CI, confidence interval; PHQ-9, 9-item patient health questionnaire; PSS-10, 10-item perceived stress scale; PROMIS, patient-reported outcomes measurement information system.

**Figure 1.**
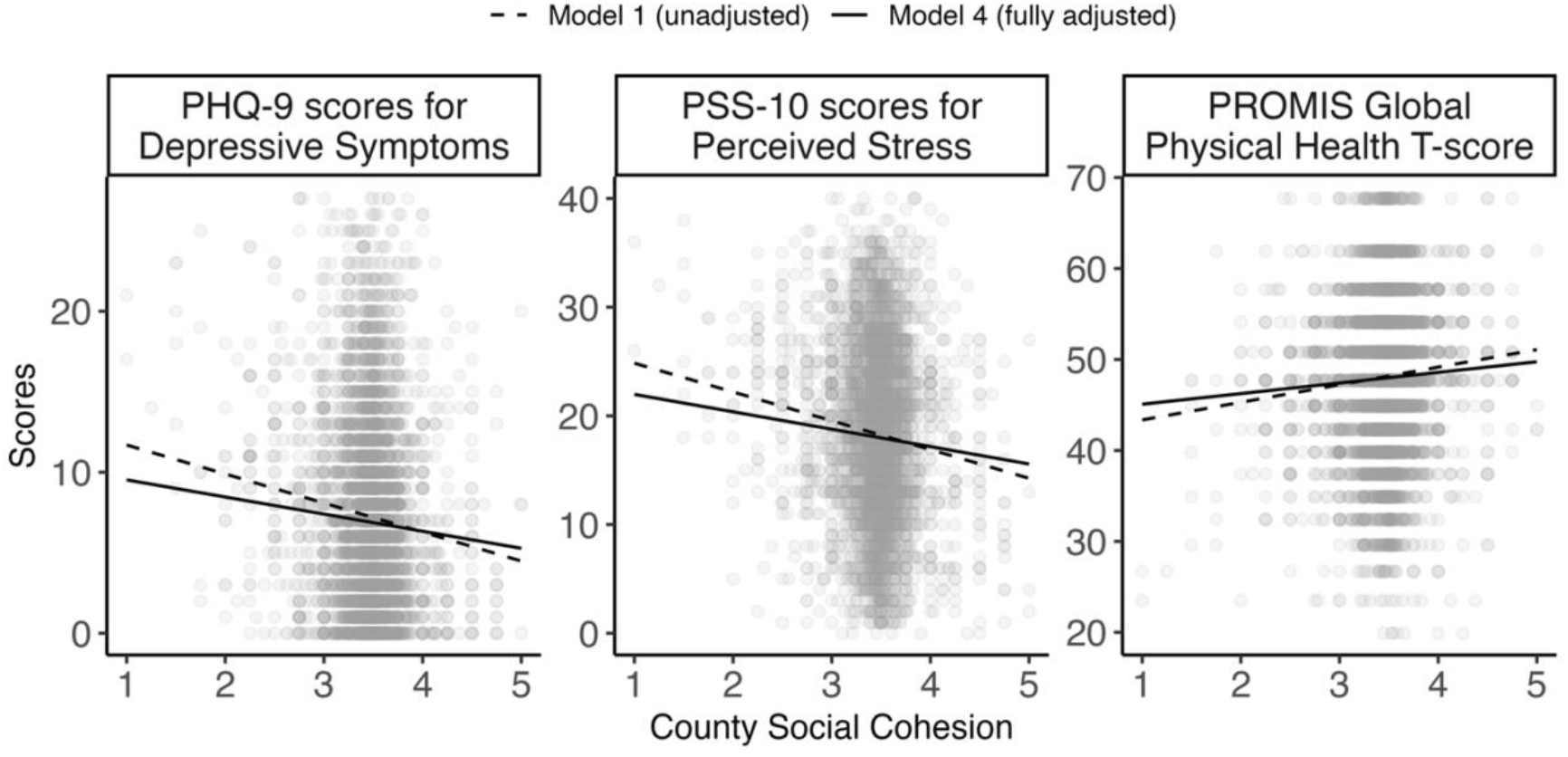
Unadjusted and fully adjusted associations between county-level social cohesion and (i) PHQ-9 scores for depressive symptoms, (ii) PSS-10 scores for perceived stress, and (iii) PROMIS Global Physical Health T-score among LGBTQIA+ adults in The PRIDE Study (no. individuals = 3752, no. counties = 778). Model 1 (unadjusted model) is represented by the dashed line and only included county-level social cohesion. Model 4 (fully adjusted model) is represented by the solid line and added mean centered age and its quadratic term, ethnoracial group, education level, gender modality, household income, proportion ≥25 years old with high school diploma (or equivalency) and proportion living below federal poverty level. Each point in the scatterplot represents an individual’s score on a mental or physical health outcome plotted against their county-level social cohesion score. PHQ-9, 9-item patient health questionnaire; PSS-10, 10-item perceived stress scale; PROMIS, patient-reported outcomes measurement information system.

**Figure 2.**
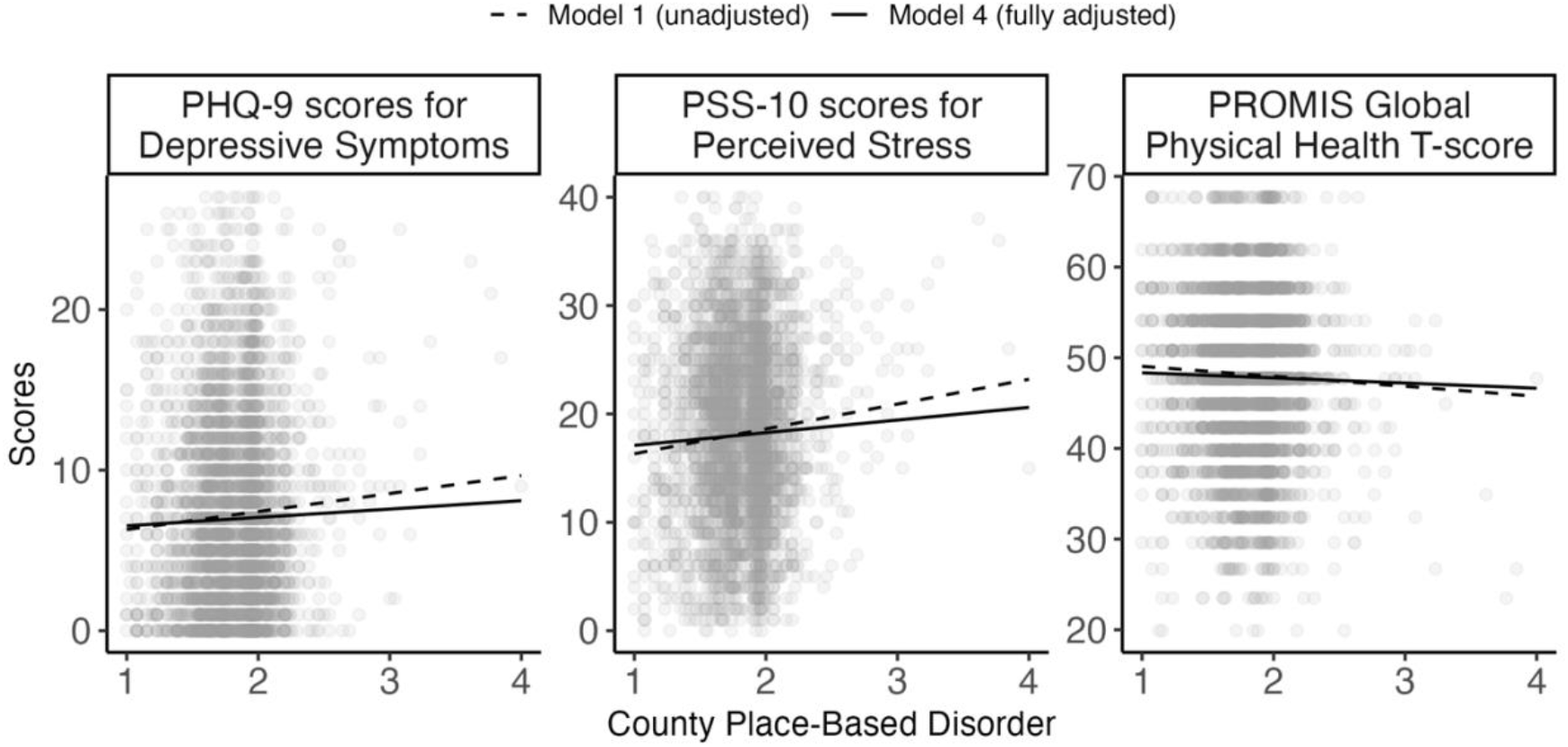
Unadjusted and fully adjusted associations between county-level place-based disorder and (i) PHQ-9 scores for depressive symptoms, (ii) PSS-10 scores for perceived stress, and (iii) PROMIS Global Physical Health T-score among LGBTQIA+ adults in The PRIDE Study (no. individuals = 3752, no. counties = 778). Model 1 (unadjusted model) only included county-level place-based disorder. Model 4 (fully adjusted model) added mean centered age and its quadratic term, ethnoracial group, education level, gender modality, household income, proportion ≥25 years old with high school diploma (or equivalency) and proportion living below federal poverty level. Each point in the scatterplot represents an individual’s score on a mental or physical health outcome plotted against their county-level disorder score. PHQ-9, 9-item patient health questionnaire; PSS-10, 10-item perceived stress scale; PROMIS, patient-reported outcomes measurement information system.

There was no evidence that the relationship between area-level characteristics and individual health outcomes differed by ethnoracial group (**Table S5**). Similarly, we did not observe effect modification by gender modality for associations involving social cohesion (**Table S6**). Only associations between place-based disorder with depressive symptoms (*P*=0.06) appeared stronger among transgender and gender diverse participants than cisgender participants. Among transgender and gender diverse participants, 1-unit higher score in place-based disorder was associated with a PHQ-9 score that is 1.11 points higher (95% CI 0.22, 1.97). These associations were not observed for cisgender participants (B=-0.10, 95% CI -1.00, 0.81).

In sensitivity analysis using unconditional empirical Bayes estimates, we observed the effects of shrinkage such that variability of the unconditional empirical Bayes estimates were smaller than for the crude means for both area-level measures (**Figure S4**). In models that regressed empirical Bayes estimates of social cohesion to each outcome, there were consistent evidence that higher levels of social cohesion was associated with lower PHQ-9 scores, lower PSS-10 scores, and higher GPH T-scores; associations were also larger in magnitude (**Table S7**). For models with empirical Bayes estimates for place-based disorder, we no longer observed a significant association with perceived stress, albiet the direction and magnitude were similar to models using the crude means.

## DISCUSSION

In this study of LGBTQIA+ adults in the US, we found that place-based disorder and lack of social social cohesion were associated with some poorer physical and mental health outcomes. This finding is consistent with the extant literature on LGBTQIA+ study populations.^17–19,21,44^ Higher place-based disorder was associated with elevated perceived stress, while higher social cohesion was associated with lower levels of depressive symptoms and perceived stress as well as better physical health. Although we did not observe effect modification by ethnoracial group, there was some evidence suggesting that gender modality modified the associations between place-based disorder and depressive symptoms with stronger associations among transgender and gender diverse participants compared to cisgender participants. These findings support our hypothesis that characteristics of the local environment may shape the health of LGBTQIA+ adults.

Area-level conditions may influence the health of LGBTQIA+ adults through multiple pathways. Adverse physical and social environments – such as those marked by disorder, neglect, or social disorganization – can elevate exposure to chronic stress and reduce perceptions of safety, contributing to poorer health.^26^ For LGBTQIA+ individuals, who already face minority stress in the form of stigma, discrimination, and social exclusion, the added burden of place-based disorder may exacerbate psychological distress.^38^ In contrast, social cohesion was shown to be potentially protective among LGBTQIA+ participants in our study. Higher social cohesion indicates greater levels of mutual trust, community belonging, and collective efficacy.^24,25^ which are factors that may enhance healthier behaviors, promote resilience, and buffer against the health-related impacts of minority stress among LGBTQIA+ individuals. A prior study has indicated that higher perception of social cohesion was associated lower odds of alcohol use before and during sex and condomless anal sex among Black sexual minority men.^17^ Another study suggests that it may also buffer against the impact of discrimination among sexual minority individuals.^44^ Thus, efforts to build levels of trust and reciprocity while addressing place-based disorder may contribute to better overall health for LGBTQIA+ residents.

Notably, our results indicate that, while the positive direction of observed association between place-based disorder and depressive symptoms is consistent with prior evidence in other non-LGBTQIA+ study populations,^38,45^ a high degree of uncertainty around the estimate remains. One possible explanation is that LGBTQIA+ communities may be forming and accessing mental health support through online spaces^46^ rather than within their immediate physical surroundings. Telehealth efforts have increased mental health care access for LGBTQIA+ individuals, and these approaches have grown since the Covid-19 pandemic.^47^ Thus, measures related to the physical environment may play less of role in shaping the mental health of LGBTQIA+ individuals. A previous study in sexual minority women indicated that census tract-level deprivation had little to no association with mental health.^20^ Similarly, another study in Black sexual minority men and transgender women showed that GPS-derived indicators of disorder were minimally associated with alcohol use.^22^

The influence of place-based disorder on health varied more distinctly by gender modality than by ethnoracial group in our study, possibly reflecting differences in how LGBTQIA+ subgroups navigate area-level contexts. While all ethnoracial minority participants contend with the structural impacts of white supremacy,^39^ the place-based impact of such a system shaped by residential segregation and discriminatory housing policies (*i*.*e*., redlining) has disproportionately affected specific ethnoracial groups, particularly Black communities.^28,29^ These structural factors may influence place-based disorder and social cohesion in different ways across ethnoracial minority populations. Our study found some evidence that levels of place-based disorder may be higher in counties with Black or African American LGBTQIA+ residents. However, due to limited sample sizes among ethnoracial minority participants, we were unable to further stratify ethnoracial categories to examine variation across subgroups. This limitation may have masked statistical interaction between ethnoracial groups and place-based disorder and social cohesion, despite prior evidence suggesting that perceptions of better area-level conditions may differentially shape the health of Black and Latino sexual minority residents.^21^

We observed potential effect modification by gender modality, with stronger associations between place-based disorder and depressive symptoms among transgender and gender diverse participants compared to cisgender peers. One explanation may be that transgender and gender diverse people are more likely to experience compounding structural vulnerabilities, including economic marginalization, housing instability, and gender-based employment discrimination, which may heighten sensitivity to area-level stressors or influence selection processes that sort transgender and gender diverse participants into more disadvantaged places of residence. Transgender and gender diverse individuals are more likely to face unemployment, poorer job quality, greater minority stress, and employment discrimination than their cisgender counterparts.^9,48,49^ These social inequities may contribute to less financial resources that would allow transgender and gender diverse participants to live in areas with greater access to supportive resources (*e*.*g*., parks, community centers, and public transportation) that may ultimately impact their mental health.

This study has several limitations. First, we used counties as the administrative unit, which captures only one particular type of spatial or social dimensions of place-based disorder and social cohesion.^26^ These constructs are, in part, rooted in perceptions of the built environment and interpersonal interactions, which are more proximal in nature and may be better represented by smaller administrative units such as census tracts. This limitation is particularly salient in rural areas, where counties span larger geographic areas and may mask meaningful heterogeneity in area-level conditions. However, our ability to conduct sensitivity analyses using smaller units was limited by the fact that over 90% of census tracts in the data included only one resident. Second, the use of crude means in the study may have introduced measurement error. Sensitivity analyses suggest high shrinkage of area-levels measures, which may be a result of poor agreement between residents in the same county or within-county small sample sizes.^43^ To our knowledge, the econometric properties of place-based scales have not been extensively evaluated in LGBTQIA+ populations. Given how LGBTQIA+ communities may interact differently with their physical and social environment, research is needed to understand how different aggregation methods of survey responses from area residents may impact the relationship between area-level characterestics and health in LGBTQIA+ communities.

Third, reverse causality may be a concern. Individuals with poorer physical or mental health may be more likely to experience downward social or economic mobility, leading them to reside in areas with higher levels of place-based disorder and lower levels of social cohesion.^26^ Fourth, evolving trends in the spatial configuration of LGBTQIA+ communities, such as the dissolution or transformation of “gayborhoods,” may influence how LGBTQIA+ individuals experience their physical and social environment.^50^ Since our study did not account for residential duration or mobility, we could not assess how long-term exposure or changes in the residential composition may shape health outcomes.

Fifth, among participants with linked health data from the annual questionnaire, excluded participants had higher depressive symptoms and perceived stress, and lower education and household income compared to included participants. This may introduced selection bias if lower-income LGBTQIA+ adults with depression living in disadvantaged areas were less likely to participate. Finally, particpants included in the study were derived from a convenience sample that is predominately white with higher education levels. Despite nationwide recruitment through a variety of community-based approaches, the fewer number of LGBTQIA+ participants from ethnoracial minority groups or with lower education levels limits the generalizability of our findings primarily to white, college-educated LGBTQIA+ adults.

Aligned with the *Healthy People 2030* goals, our findings add to a growing body of evidence that highlights the relevance of area-level characteristics in understanding the health of LGBTQIA+ populations. These results underscore the need for policies and interventions that aim at preventing disinvestment from LGBTQIA+ communities, enhancing social cohesion, and promoting equity in the physical environment. Place-based research is thus critical for understanding how broader structural conditions shape health inequities beyond individual-level exposures. Future research should include more granular geographic data, longitudinal designs, and residential histories to better understand how LGBTQIA+ people’s environments shape their health outcomes.

## Data Availability

The datasets generated during and/or analyzed during the current study are not publicly available due to ethical restrictions related to sensitive participant information but are available from the corresponding author on reasonable request. Researchers interested in The PRIDE Study data may submit a brief application which is reviewed by both a Research Advisory Committee (composed of scientists) and Participant Advisory Committee (composed of participants) to affirm appropriate data use. Details about the Ancillary Study process are available at www.pridestudy.org/collaborate or by contacting or 855-421-9991 (toll-free).

