## Supplemental for "Place-based disorder, social cohesion, and physical and mental health outcomes of LGBTQIA+ adults in the US"

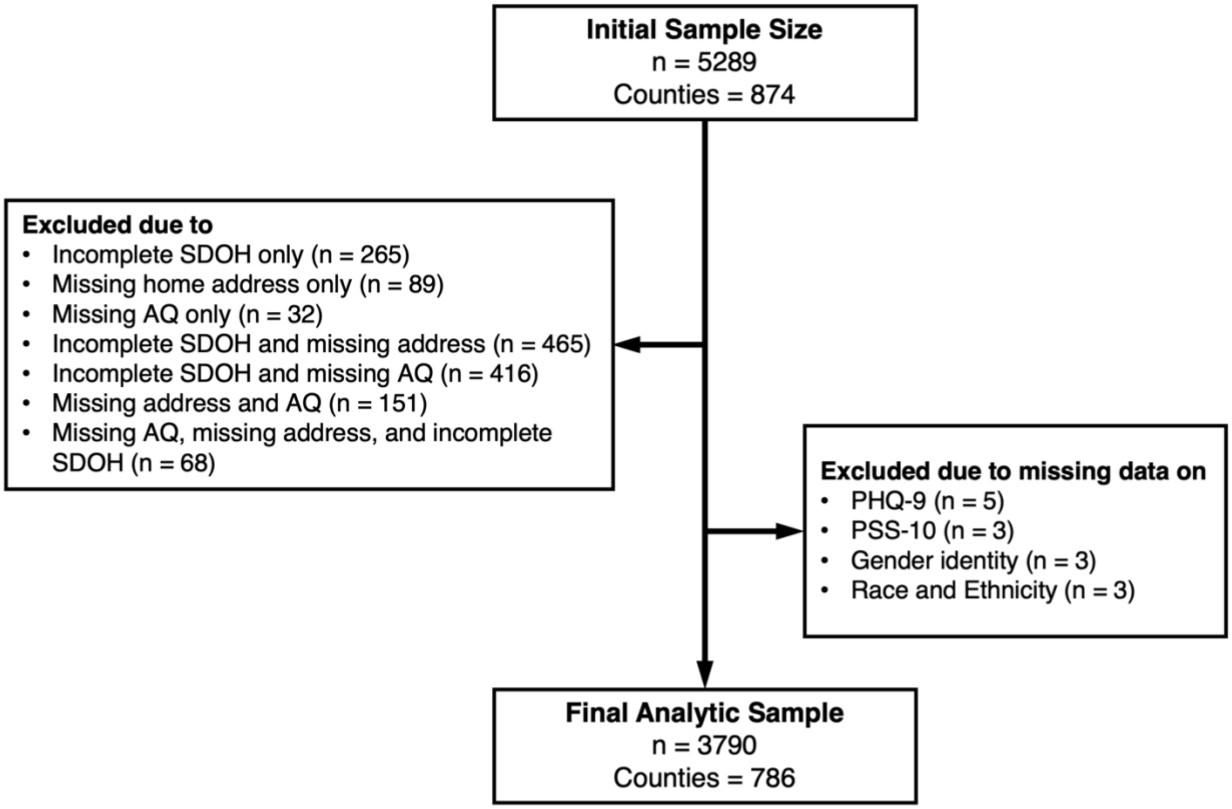

**Figure S1**. Flow chart describing the sample inclusion criteria of the analytic sample from The PRIDE Study.

AQ, annual questionnaire; PHQ-9, 9-item patient health questionnaire; PSS-10, 10-item perceived stress scale; PROMIS, patient-reported outcomes measurement information system; SDOH, social determinants of health ancillary questionnaire.

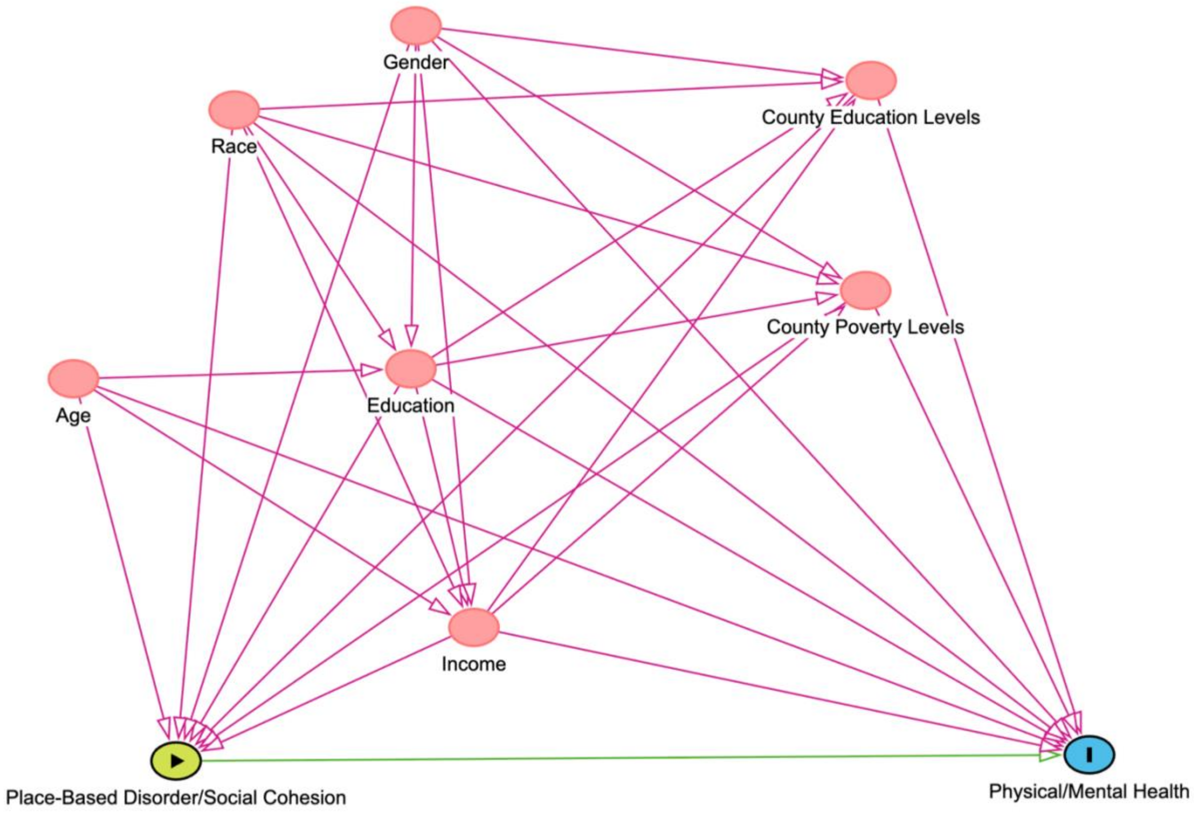

**Figure S2.** Directed acyclic graph identifying potential confounders for regression adjustment based on published literature.

Source for software: Textor J, van der Zander B, Gilthorpe MK, Liskiewicz M, Ellison GTH. Robust causal inference using directed acyclic graphs: the R package 'dagitty'. Int J Epidemiol 2016; 45(6):1887-1894.

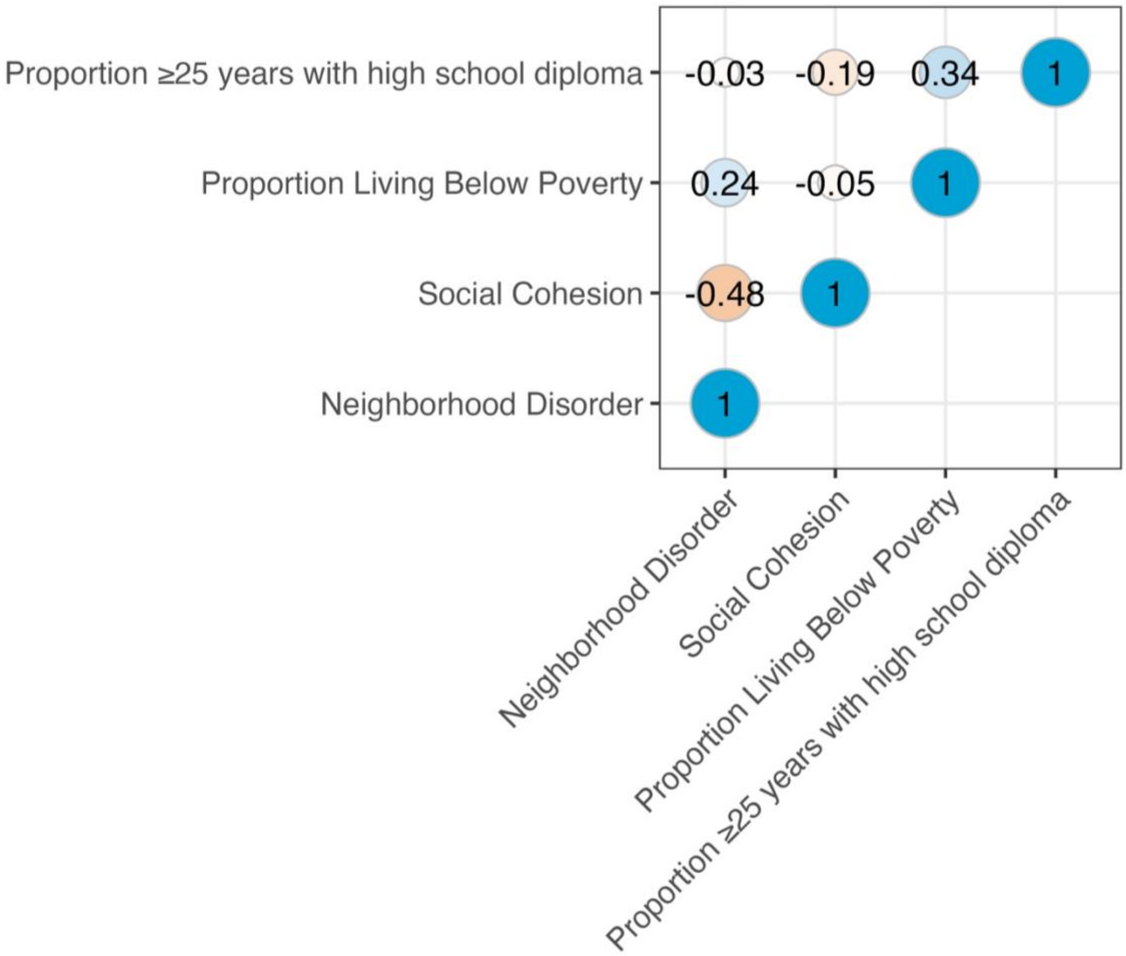

**Figure S3.** Correlation between county-level area characteristics and socioeconomic conditions.

The correlation coefficient measures the strength and directional of the linear association between two variables. It ranges from −1 to 1, with zero indicating no correlation. Blue indicates that variables are positive correlated with one another, and orange indicates variables are negatively correlated.

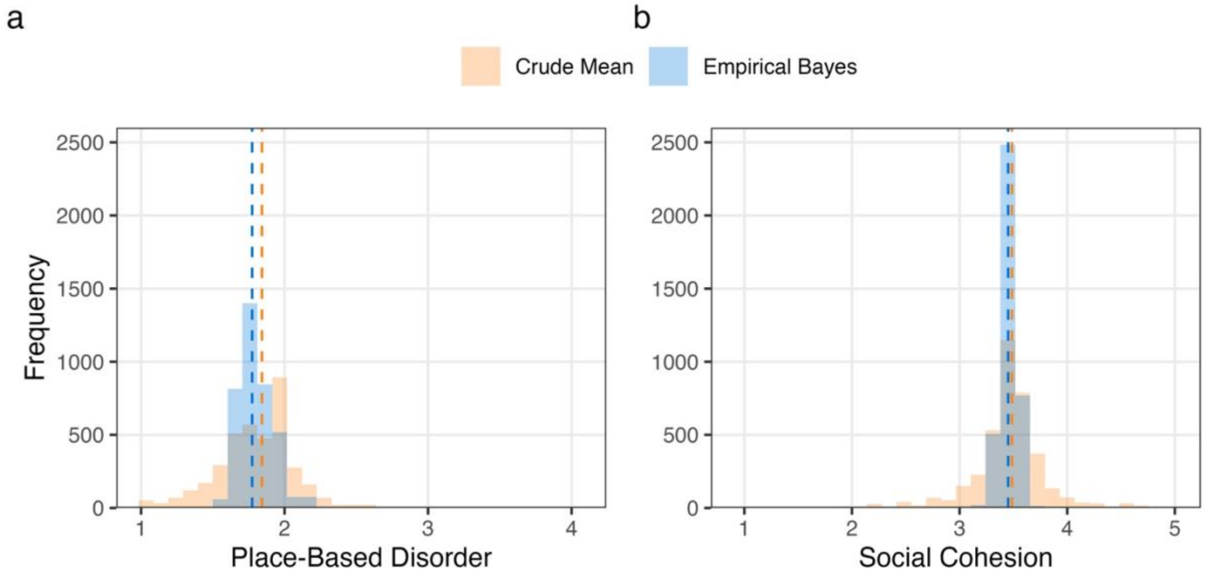

**Figure S4.** Distribution of crude means and unconditional empirical Bayes estimates for (a) place-based disorder and (b) social cohesion for across counties in the study population (no. counties = 778). Vertical dashed lines indicate the median for the crude means and empirical Bayes estimates.

**Table S1.** Comparison between included and excluded participants with linked annual health questionnaire data.

| | Included | Excluded | |
|---|---|---|---|
| | n = 3790 | n = 612 | *P* |
| PHQ-9 score for Depressive Symptoms, mean (SD) | 7.3 (6.0) | 8.1 (6.2) | 0.003 |
| PSS-10 score for Perceived Stress, mean (SD) | 18.5 (8.2) | 19.7 (8.0) | 0.008 |
| PROMIS Global Physical Health T-score, mean (SD) | 47.9 (8.5) | 47.7 (8.9) | 0.64 |
| Age, median (IQR), y | 37.2 (29.4-54.0) | 32.7 (26.0-43.7) | <0.001 |
| Gender Identity, No. (%) | | | <0.001 |
| Cisgender man | 953 (25.1) | 112 (18.3) | |
| Cisgender woman | 1102 (29.1) | 185 (30.2) | |
| Non-binary | 896 (23.6) | 147 (24.0) | |
| Transgender man | 464 (12.2) | 90 (14.7) | |
| Transgender woman | 213 (5.6) | 38 (6.2) | |
| Another gender identity | 162 (4.3) | 36 (5.9) | |
| Missing | 0 (0.0) | 4 (0.7) | |
| Sexual Orientation, No. (%) | | | 0.04 |
| Asexual/Demisexual/Gray-Ace | 319 (8.4) | 64 (10.5) | |
| Bisexual/Pansexual | 891 (23.5) | 171 (27.9) | |
| Gay/Lesbian | 1593 (42.0) | 224 (36.6) | |
| Queer | 910 (24.0) | 142 (23.2) | |
| Straight/Heterosexual | 54 (1.4) | 6 (1.0) | |
| Another sexual orientation | 23 (0.6) | 5 (0.8) | |
| Ethnoracial Identity, No. (%) | | | |
| American Indian or Alaska Native | 109 (2.9) | 14 (2.3) | 0.50 |
| Asian | 171 (4.5) | 30 (4.9) | 0.73 |
| Black or African American | 158 (4.2) | 28 (4.6) | 0.71 |
| Hispanic or Latino | 246 (6.5) | 44 (7.2) | 0.56 |
| Middle Eastern or North African | 48 (1.3) | 7 (1.1) | 0.96 |
| Native Hawaiian or Pacific Islander | 10 (0.3) | 1 (0.2) | 0.98 |
| White | 3478 (91.8) | 558 (91.2) | 0.87 |
| More than one ethnoracial identity | 412 (10.9) | 71 (11.6) | 0.62 |
| Another ethnoracial identity | 58 (1.5) | 11 (1.8) | 0.74 |
| Education Level, No. (%) | | | <0.001 |
| High school or less | 162 (4.3) | 40 (6.5) | |
| Some college | 719 (19.0) | 147 (24.0) | |
| 4-year degree | 1252 (33.0) | 225 (36.8) | |
| Graduate degree | 1655 (43.7) | 200 (32.7) | |
| Missing | 2 (0.1) | 0 (0.0) | |
| Annual Household Income, No. (%) | | | 0.004 |

| | | |
|---|---|---|
| $0-20,000 | 488 (12.9) | 91 (14.9) |
| $20,001-50,000 | 856 (22.6) | 149 (24.3) |
| $50,001-80,000 | 698 (18.4) | 104 (17.0) |
| $80,001-100,000 | 374 (9.9) | 45 (7.4) |
| $100,001-150,000 | 621 (16.4) | 102 (16.7) |
| >$150,000 | 716 (18.9) | 105 (17.2) |
| Missing | 37 (1.0) | 16 (2.6) |

*P* based on $\chi^2$ test for categorical variables or t-test/Mann-Whitney U-test for continuous variables. IQR, interquartile range; PHQ-9, 9-item patient health questionnaire; PSS-10, 10-item perceived stress scale; PROMIS, patient-reported outcomes measurement information system; SD, standard deviation.

**Table S2.** Scale items used to measure area characteristics in the Social Determinants of Health ancillary survey.

| Area Characteristic | Scale Item |
|---|---|
| Place-based disorder | There is a lot of graffiti in my neighborhood. |
| | My neighborhood is noisy. |
| | Vandalism is common in my neighborhood. |
| | There are a lot of abandoned buildings in my neighborhood. |
| | My neighborhood is clean. |
| | People in my neighborhood take good care of their houses and apartments. |
| | There are too many people hanging around on the streets near my home. |
| | There is a lot of crime in my neighborhood. |
| | There is too much drug use in my neighborhood. |
| | There is too much alcohol use in my neighborhood. |
| | I'm always having trouble with my neighbors. |
| | In my neighborhood, people watch out for each other. |
| | My neighborhood is safe. |
| Social cohesion | People around here are willing to help their neighbors. |
| | People in my neighborhood generally get along with each other. |
| | People in my neighborhood can be trusted. |
| | People in my neighborhood share the same values. |

**Table S3.** Sample size distribution of sexual and gender minority residents by the number of counties represented.

| | Median | Range | Number of Counties | % |
|---|---|---|---|---|
| Number of participants per county | 2 | 1-114 | | |
| 1 | | | 368 | 46.8 |
| 2 | | | 119 | 15.1 |
| 3-5 | | | 147 | 18.7 |
| 6-10 | | | 80 | 1.02 |
| More than 10 | | | 72 | 9.2 |

**Table S4.** Variance components for individual health outcomes.

| | Variance | | |
|---|---|---|---|
| | Individual n = 3752 | County n = 778 | ICC |
| PHQ-9 score for Depressive Symptoms | 34.802 | 1.457 | 0.040 |
| PSS-10 score for Perceived Stress | 65.279 | 1.790 | 0.027 |
| PROMIS Global Physical Health T-score | 70.137 | 2.937 | 0.040 |

Results were based on a two-level null model with random intercept for counties. ICC, intraclass correlation coefficient; PHQ-9, 9-item patient health questionnaire; PSS-10, 10-item perceived stress scale; PROMIS, patient-reported outcomes measurement information system.

**Table S5.** Mean differences (95% CI) for each individual health outcome per 1-unit higher score in place-based disorder and social cohesion by ethnoracial group, The PRIDE Study.

| | Mean differences (*B*) in outcome (95% CI) by 1-unit higher score of area characteristic | | |
|---|---|---|---|
| | Minoritized Ethnoracial Group | White Only | Global *P* for interaction |
| | n = 687 | n = 3065 | |
| ***PHQ-9 scores for Depressive Symptoms*** | | | |
| Place-based Disorder | 0.23 (-1.31, 1.76) | 0.58 (-0.12, 1.27) | 0.68 |
| Social Cohesion | -1.09 (-2.38, 0.19) | -1.06 (-1.60, -0.52) | 0.96 |
| ***PSS-10 scores for Perceived Stress*** | | | |
| Place-based Disorder | 1.24 (-0.79, 3.37) | 1.16 (0.24, 2.07) | 0.94 |
| Social Cohesion | -1.54 (-3.24. 0.16) | -1.61 (-2.32, -0.90) | 0.94 |
| ***PROMIS Global Physical Health T-score*** | | | |
| Place-based Disorder | -0.83 (-3.08, 1.42) | -0.51 (-1.55, 0.53) | 0.79 |
| Social Cohesion | 2.36 (0.48, 4.24) | 0.97 (0.19, 1.76) | 0.18 |

All models adjusted for mean centered age and its quadratic term, gender modality, education level, household income, proportion ≥25 years old with high school diploma (or equivalent), and proportion living below federal poverty level. Global *P* for interaction was based on Likelihood Ratio test. CI, confidence interval; PHQ-9, 9-item patient health questionnaire; PSS-10, 10-item perceived stress scale; PROMIS, patient-reported outcomes measurement information system.

**Table S6.** Mean differences (95% CI) for each individual health outcome per 1-unit higher score in place-based disorder and social cohesion by gender modality, The PRIDE Study

| | Mean differences (*B*) in outcome (95% CI) by 1-unit higher score of area characteristic | | |
|---|---|---|---|
| | Cisgender | Transgender and Gender Diverse | Global *P* for interaction |
| | n = 2035 | n = 1717 | |
| ***PHQ-9 scores for Depressive Symptoms*** | | | |
| Place-based Disorder | -0.10 (-1.00, 0.81) | 1.11 (0.22, 1.97) | 0.06 |
| Social Cohesion | -0.99 (-1.69, -0.28) | -1.14 (-1.84, -0.45) | 0.75 |
| ***PSS-10 scores for Perceived Stress*** | | | |
| Place-based Disorder | 0.67 (-0.52, 1.86) | 1.64 (0.48, 2.79) | 0.25 |
| Social Cohesion | -1.59 (-2.52, -0.66) | -1.61 (-2.53, -0.69) | 0.98 |
| ***PROMIS Global Physical Health T-score*** | | | |
| Place-based Disorder | -0.05 (-1.39, 1.29) | -1.03 (-2.32, 0.26) | 0.28 |
| Social Cohesion | 1.30 (0.27, 2.33) | 1.04 (0.03, 2.06) | 0.73 |

All models adjusted for mean centered age and its quadratic term, ethnoracial group, education level, household income, proportion ≥25 years old with high school diploma (or equivalent), and proportion living below federal poverty level. Global *P* for interaction was based on Likelihood Ratio test. CI, confidence interval; PHQ-9, 9-item patient health questionnaire; PSS-10, 10-item perceived stress scale; PROMIS, patient-reported outcomes measurement information system.

**Table S7.** Association of area-level characteristics using the unconditional empirical Bayes estimates and individual-level depressive symptoms, perceived stress, and general physical health, The PRIDE Study.

| | No. Individuals | No. Counties | Place-Based Disorder | | Social Cohesion | |
|---|---|---|---|---|---|---|
| | | | *B* (95% CI) | *P* | *B* (95% CI) | *P* |
| ***PHQ-9 scores for Depressive Symptoms*** | | | | | | |
| Model 1 | 3752 | 778 | 3.58 (1.09, 6.07) | 0.005 | -12.8 (-16.1, -9.52) | <0.001 |
| Model 2 | 3752 | 778 | 1.97 (-0.28, 4.23) | 0.09 | -10.9 (-13.9, -7.90) | <0.001 |
| Model 3 | 3752 | 778 | 0.24 (-1.33, 1.81) | 0.77 | -5.87 (-8.38, -3.37) | <0.001 |
| Model 4 | 3752 | 778 | 1.04 (-0.60, 2.69) | 0.22 | -4.81 (-7.40, -2.21) | <0.001 |
| ***PSS-10 scores for Perceived Stress*** | | | | | | |
| Model 1 | 3752 | 778 | 5.97 (2.79, 9.15) | <0.001 | -17.6 (-21.9, -13.3) | <0.001 |
| Model 2 | 3752 | 778 | 2.66 (0.09, 5.22) | 0.04 | -13.8 (-17.4, -10.2) | <0.001 |
| Model 3 | 3752 | 778 | 0.92 (-1.00, 2.85) | 0.35 | -7.88 (-11.0, -4.73) | <0.001 |
| Model 4 | 3752 | 778 | 1.50 (-0.57, 3.56) | 0.15 | -7.53 (-10.8, -4.24) | <0.001 |
| ***PROMIS Global Physical Health T-score*** | | | | | | |
| Model 1 | 3752 | 778 | -3.31 (-6.77, 0.16) | 0.06 | 17.1 (12.5, 21.7) | <0.001 |
| Model 2 | 3752 | 778 | -2.58 (-5.99, 0.83) | 0.14 | 16.3 (11.8, 20.8) | <0.001 |
| Model 3 | 3752 | 778 | -0.30 (-2.96, 2.37) | 0.83 | 8.73 (4.83, 12.6) | <0.001 |
| Model 4 | 3752 | 778 | -1.28 (-3.99, 1.43) | 0.36 | 7.81 (3.82, 11.8) | <0.001 |

Models 1 included each area characteristic in a separate model. Model 2 added mean centered age and its quadratic term. Model 3 added ethnoracial group, education level, gender modality, and household income. Model 4 added proportion ≥25 years old with high school diploma (or equivalency) and proportion living below federal poverty level. CI, confidence interval; PHQ-9, 9-item patient health questionnaire; PSS-10, 10-item perceived stress scale; PROMIS, patient-reported outcomes measurement information system.
